# “Look straight ahead” – A new test to diagnose spatial neglect by computed tomography

**DOI:** 10.1101/2022.08.22.22278887

**Authors:** Joel Coelho Marques, Jens Hanke, Caroline Schell, Frank Andres, Hans-Otto Karnath

**Affiliations:** Department of Neurology and Early Rehabilitation, Kreiskliniken Reutlingen GmbH, Reutlingen, Germany; Diagnostic and Interventional Radiology, Kreiskliniken Reutlingen GmbH, Reutlingen, Germany; Center of Neurology, Division of Neuropsychology, Hertie-Institute for Clinical Brain Research, University of Tübingen, Tübingen, Germany

**Author notes:** Correspondence should be addressed to: Hans-Otto Karnath, Centre of Neurology, University of Tübingen, 72076 Tübingen, Germany.

**Keywords:** Spatial neglect, Conjugate eye deviation (CED), Attention, Neuroimaging, Stroke, Human

## Abstract

Spatial neglect is the dominant behavioral disorder after right hemisphere brain lesions. Reliable diagnosis by formal neuropsychological testing is often achieved only later during hospitalization, leading to delays in targeted therapies. We propose a way to diagnose spatial neglect right at admission. We measured the conjugated eye deviation (CED) on the initial computed tomography (CT) scans, in combination with the verbal instruction “Please look straight ahead” during the scan. The command was implemented in the scanner program and automatically played before a cranial CT started. This prospective study included a total 46 consecutive subjects (16 patients with first ever right brain damage and no spatial neglect, 12 patients with first ever right brain damage and spatial neglect, and 18 healthy controls). The right brain damaged groups were submitted to paper pencil tests to access the diagnosis of a spatial neglect after radiological confirmation of the brain damage during the initial phase of their hospitalisation. This procedure allowed us to define a cut-off value of 14.1 degrees of CED to the ipsilesional side to differentiate right hemispheric stroke patients with versus without spatial neglect with a confidence interval of 99%. This simple addition to a radiological routine procedure provides a new tool to help diagnose spatial neglect at the earliest stage possible and thus offers the possibility of providing patients with optimized rehabilitative therapy from a very early stage on.

## Introduction

Despite declining incidence and mortality [1], cerebrovascular disease stays of upmost importance as a health indicator, especially in terms of resulting disability, associated healthcare, and nursing costs [2,3]. Along with hemiparesis, the two most dominant clinical symptoms of stroke are aphasia after a lesion of the human left hemisphere and spatial neglect after right hemisphere damage [4]. Early diagnosis of these disorders is important for several reasons: Studies have shown that early onset rehabilitative therapy within the first 24 hours after stroke can improve the outcomes of the neurological deficits [4,5] - aphasia and spatial neglect included [5] - if the interventions are frequent and short [5]. Furthermore, spatial neglect has been shown to increase hospitalization times and slow down the recovery from additional deficits [6], which highlights the importance of an early diagnosis.

While early assessment of language disorders in awake patients is straightforward, the diagnosis of spatial neglect is not so well established in this (hyper)acute phase of admission, i.e. when paper-and-pencil testing is often not yet feasible. Two early clinical signs in neglect patients are the spontaneous and sustained deviation of the eyes (conjugate eye deviation [CED]) and of the head toward the ipsilesional side [7–9]. Becker and Karnath [7] observed that the horizontal eye-in-head deviation is specifically associated with spatial neglect rather than with brain damage per se and that it can be detected already in clinical imaging scans taken at admission.

Using the latter observation for a clinical test of spatial neglect seemed obvious. However, the retrospective analysis of routine clinical scans by Becker and Karnath [7], i.e., scans obtained without further modifications of the typical neuroradiological imaging procedure, did not allow a control of the patients’ eye-in-head position; they were free to direct their eyes in any direction.

In the present prospective study, we tried to maximize the discrepancy between horizontal eye-in-head deviation between stroke patients with and without spatial neglect by giving the simple verbal command “Please look straight ahead” during the scan. While this instruction can easily be followed by stroke patients without neglect, it will not help patients with neglect to overcome their tonic horizontal eye-in-head deviation. This simple verbal instruction could therefore serve to maximize the discrepancy between these two groups and might allow to build a formal cut-off for differentiation between them.

## Methods

### Subjects

Neurological patients with the suspicion of an acute stroke consecutively admitted between February 2018 and March 2020 to the Department of Neurology in Reutlingen were screened for a first ever right-hemisphere stroke. Patients with tumors, patients in whom MRI or CT scans revealed no obvious lesions, as well as patients with disturbed awareness at admission were not included. Patients who were not able to perform the paper/pencil tasks and patients who underwent any type of revascularization therapy were also excluded (see below). We also did not include patients with left-sided stroke to exclude any conflicts between the here newly applied (verbal) procedure in the scanner (see below) and possible disturbances of language processing [10]. A group of control subjects consisted of 18 subjects in whom CT imaging had been conducted due to headache, but no pathological findings had been revealed. Clinical and demographic data of all subjects are presented in Table 1.

**Table 1.** Clinical and demographic data of all subjects included with acute first ever right hemispheric stroke.

|  |  | **Patients with spatial neglect** | **Patients without spatial neglect** |
| --- | --- | --- | --- |
| **Gender** |  |  |  |
| - Male |  | 5 | 8 |
| - Female |  | 7 | 8 |
| **Age (years)** | Mean (SD) | 74,9 (10,7) | 79,1 (9,3) |
| **Hemiparesis** | Number of Patients (%) | 11 (91,7) | 14 (87,5) |
| **Hypesthesia** | Number of Patients (%) | 9 (75) | 7 (43,75) |
| **Visual field defect** | Number of Patients (%) | 1 (8,3) | 0 (0) |
| **NIH-SS score at admission** | Mean (SD) | 9,9 (4,4) | 3,4 (2,6) |
| **Letter cancelation (CoC)** | Mean (SD) | 0,353 (0,246) | 0,003 (0,010) |
| **Bells test (CoC)** | Mean (SD) | 0,484 (0,224) | 0,011 (0,010) |
| **Albert's test (CoC)** | Mean (SD) | 0,238 (0,332) | 0 (0) |
| **Copy task (n omitted)** | Mean (SD) | 4,4 (1,7) | 0 (0) |

### Neuroimaging and procedure

Structural imaging was acquired by computed tomography (CT) as part of the clinical routine procedure carried out for all acute stroke patients in the Department of Neurology in Reutlingen at admission (Siemens Somatom Definition AS 64 or AS 40). During the scan, the command “please look straight ahead” was given twice. The verbal command was recorded and its playback integrated in the automatic program of the CT scanner to assure 100% reproducibility. The first time the verbal command was given while the patient was positioned on the CT-table; the second time immediately before the scan started. It was assured that there were no physical landmarks on the CT scanners that would help the patients find “straight ahead”. The scans had either 40 or 64 slices with a resolution of 0.6mm on acquisition. After acquisition the slices were processed with 4mm and 0.75mm distance between slices. The initial scans were used for the measurement of eye deviation in the present study (see below). They were performed on average 19.3 hours (SD 22.3) after stroke-onset. The average time to first scan is longer compared to the overall departmental average because we had to exclude all patients who underwent any type of revascularization therapy. This exclusion was necessary to avoid false-negative results [11-13], i.e., patients who had spatial neglect during the initial CT scan at admission, but no longer after successful revascularization, i.e. in the phase when the behavioral examination for spatial neglect became possible and was performed (see below). Stroke diagnosis was based on the initial scans in combination with follow-up imaging (mostly MRI), in those cases in which lesions were not yet visible on the initial scans.

### Analysis of brain scans

#### Measurement of eye deviation

We based the evaluation of the eye-in-head orientation on the technique by Simon and co-workers [11]. Horizontal deviation of eye-in-head position was defined by the angle formed between the intersection of the ocular axis and the “line of best fit” through the midline structures of the head (Fig. 1). Angles were measured with the angle measurement tool from the radiological imaging program IDS7, Sectra PACS (version 19.3.3, November 2017, Manufacturer Sectra AB, Linköping, Sweden). During the angle measurement the scans were in DICOM format and with no modification to the admission specifications. The angles of the left and the right eye were averaged for each individual to give the final deviation angle of the eyes. Horizontal deviations towards the ipsilesional side were coded as positive values; deviations to the contralesional side as negative values. For the non-brain damaged control group rightward deviations were coded as positive values; leftward deviations as negative values.

**Figure 1.**
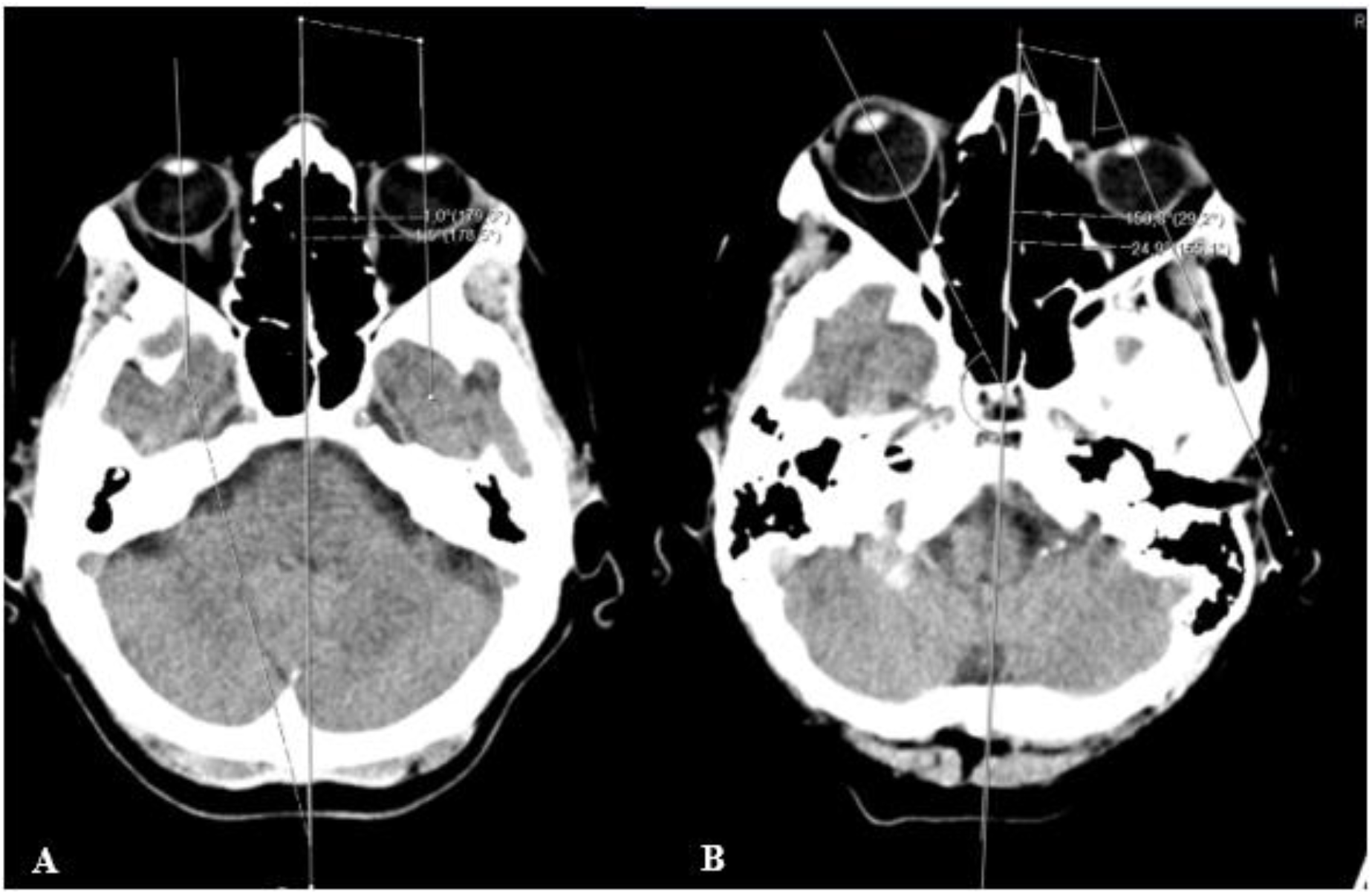
Measurement of the conjugated eye deviation (CED) on a healthy control subject with almost no deviation from midline (A) and on a patient with spatial neglect (B). A line was drawn through the middle section as a,, line of best fit“, another two lines were drawn through the ocular axis [11]. The numbers correspond to the measured angles of intersection between the midline and the line of the respective ocular axis.

#### Analysis of brain lesions

Lesion boundaries were delineated in a semi-automated way after file conversion from DICOM to NII using the Clusterize algorithm on the SPM Clusterize toolbox [12] on SPM 12 (www.fil.ion.ucl.ac.uk/spm). Normalization of CT or MR scans to MNI space with 1×1×1 mm resolution was performed by using the Clinical Toolbox [13] under SPM12, and by registering lesions to its age-specific CT templates oriented in MNI space [13]. Delineation of lesion borders and quality of normalization were verified by consensus of always two experienced investigators (one of them H.-O.K.). An overlap of the normalized lesions of the two brain damaged groups is shown in Fig. 2. The average lesion size in the sample of patients without spacial neglect was 10,4 cm^3^ (SD 19,9 cm^3^) and in the sample with spatial neglect 72,2 cm^3^ (SD 59,0 cm^3^).

**Figure 2.**
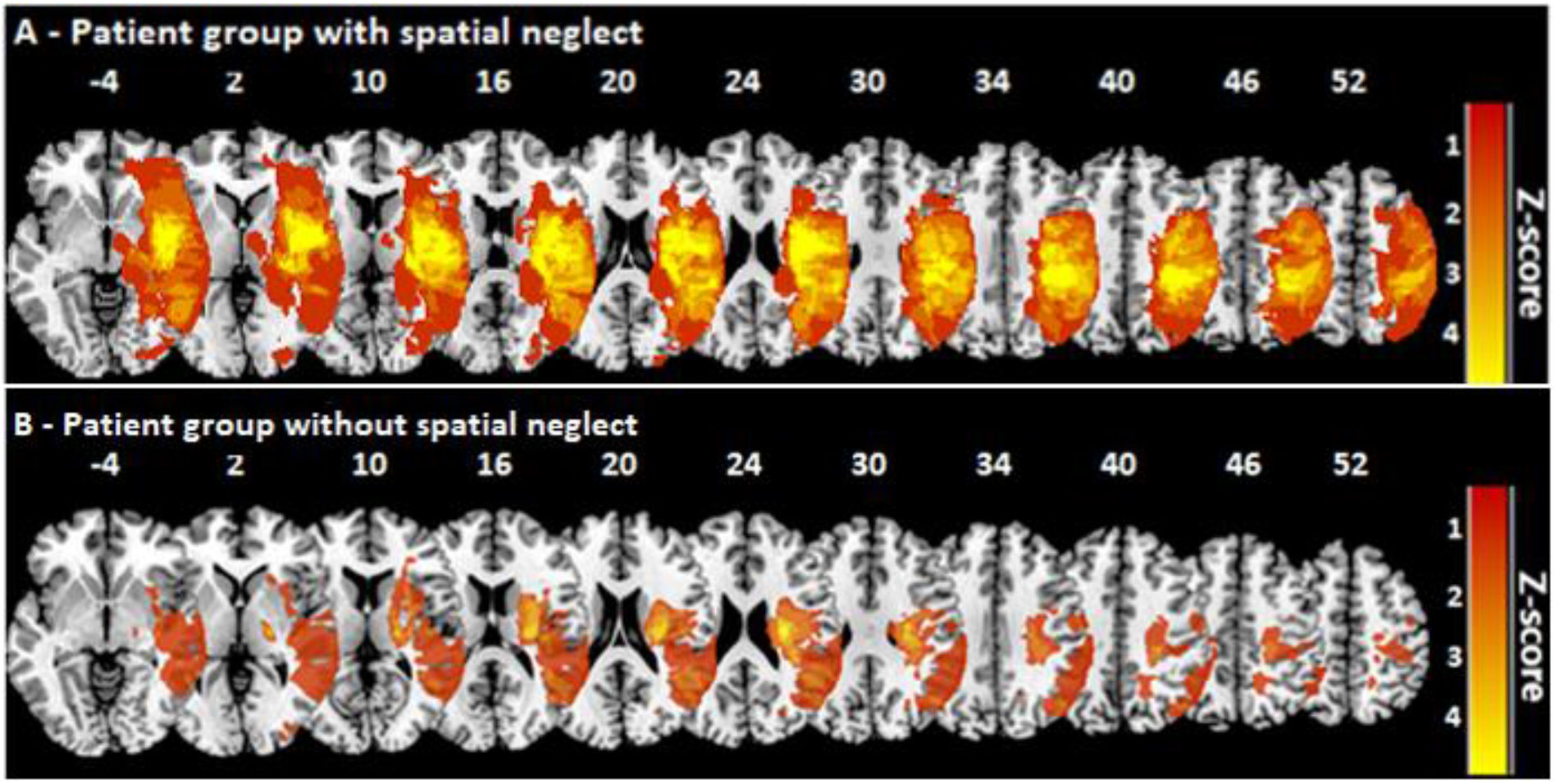
Overlap of the normalized lesions of the two right brain damaged patient groups. The lesion maps were superimposed on the single-subject T1 MNI152 template. For each voxel, the number of patients with a lesion at that location is color coded. The vertical z coordinate for each slice of standardized MNI space is given.

#### Behavioral examination

Right at admission the National Institutes of Health Stroke Scale (NIH-SS) [14] was performed for each patient by the respective neurologist on duty at the Department of Neurology in Reutlingen. Visual field defects were examined by the common neurological confrontation technique. Beyond and in parallel to the procedure used by Becker and Karnath [7], the following neuropsychological tests were performed: Letter Cancellation Task [15], the Bells Test [16], the Albert’s test [17], and a copying task [18]. This neuropsychological examination was carried out by J.C.M. and took place after the radiological confirmation of right brain damage and after assessment of the inclusion and exclusion criteria, on average 76.5 hours (SD 107.4) after initial image acquisition. The tests were presented on a horizontally oriented 21 × 29.7 cm sheet of paper which was fixed at the center of the patient’s sagittal midline. In the Letter Cancellation task, 60 target letters ‘A’ are distributed among other distractor letters [15]. The Bells test requires identifying 35 bell icons distributed all over the sheet between other symbols [16]. In these two cancellation tasks, patients were asked to cancel all of the targets, ‘A’ letters or bells respectively [15,16]. The Albert’s test consists of seven columns of 36 black lines; three on the left side and three on the right side of the horizontally orientated sheet of paper [17]. Patients had to cancel all lines. For the Letter and Bells Cancellation tasks as well as the Albert’s task, we calculated the Center of Cancellation (CoC) using the procedure and cut-off scores for diagnosing spatial neglect by Rorden and Karnath [19]. The CoC is a sensitive measure capturing both the number of omissions, as well their location [19]. In the copying task, patients were asked to copy a complex multi-object scene consisting of four figures (a fence, a car, a house, and a tree), two in each half of the horizontally oriented sheet of paper [18]. Omission of at least one of the contralateral features of each figure was scored as 1, and omission of each whole figure was scored as 2 [18]. One additional point was given when contralateral located figures were drawn on the ipsilesional side of the paper sheet [18]. The maximum score was 8 [18]. A score higher than 1 (i.e. > 12.5% omissions) was taken to indicate neglect [18]. The maximum duration of each test was not fixed in advance but depended on the patient being satisfied with his performance and confirming this twice. Following the procedure used by Becker and Karnath [7], for a safe diagnosis of spatial neglect two of the four clinical tests for spatial neglect had to be positive. For a safe exclusion of spatial neglect none of the four clinical neglect tests had to be positive. This led to the exclusion of 4 subjects in which only one of the four tests was positive. Data of all subjects are presented in Table 1.

## Results

In the sample of 28 stroke patients with right brain damage, 12 patients showed spatial neglect (cf. Table 1). Figure 3 illustrates the degree of horizontal eye deviation in the two groups of brain damaged patients as well as the non-brain damaged subjects. An ANOVA (SPSS software; vers. 24; SPSS Inc., Chicago, IL, U.S.A.) with factor subject group (spatial neglect, no spatial neglect, non-brain damaged controls) revealed a significant result (F(2,43)=33.961, p <0.001; η_p_^2^=0.612). Post-hoc comparisons revealed that the degree of eye deviation towards the ipsilesional side was significantly larger in the patients with spatial neglect than the brain damaged subjects without the disorder (mean 4.5 [SD 4.1]; t(28)=-6.731, p<0.001) and than the non-brain damaged subjects (mean 1.4 [SD 3.3]; t(26)= -5.349, p<0.001).

**Figure 3.**
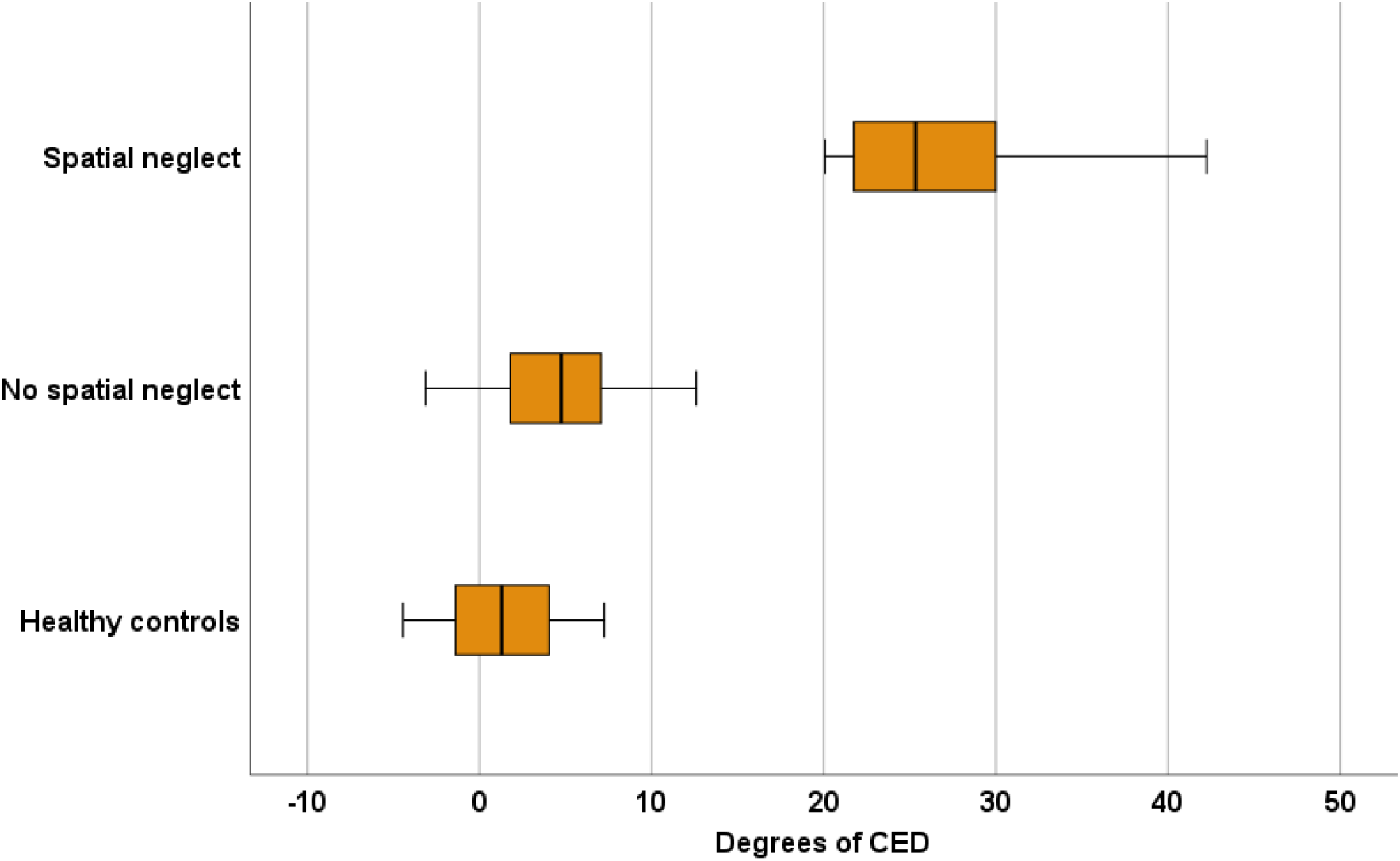
Degree of the horizontal eye deviation of the three subject groups. Positive values indicate horizontal deviation towards the ipsilesional side; negative values horizontal deviation towards the contralesional side. The CED of each patient was calculated through the average of the deviation of both eyes. The boxplot shows the median and quartile distribution of the CED on the different subject groups.

The acute right-sided stroke patients with spatial neglect had a mean CED score of 23.55 (SD 12.80). To assess the sensitivity of the CED score, we used 2.326 standard deviations to create a cut-off threshold of 14.06 – this value corresponds to p < 0.01 for a one-tailed test. To validate the sensitivity of this measure, we applied this threshold to the patients who were classified as having spatial neglect. The CED threshold was able to correctly detect 11 out of the 12 individuals with neglect. On the other hand, if this threshold was applied to the 16 stroke patients without spatial neglect (the population used to define our threshold), a total of 0 individuals were falsely classified as having spatial neglect. Therefore, out of the 28 individuals, our binary CED cut-off score agreed with the independent traditional scoring method applied to three paper-and-pencil neglect tests in 96.4% of the cases. We also computed the Receiver Operating Characteristics (ROC) value known as the ‘Area Under the Curve’ (AUC), using the formula described by Obuchowski [20]. We found that the CED score was a high accuracy predictor of spatial neglect as defined by the traditional paper-and-pencil neglect tests; the AUC was 0.9167 (i.e. was close to a ‘perfect’ AUC of 1.0).

## Discussion

Previous studies of the horizontal eye-in-head deviation after stroke, i.e. the tonic CED to the ipsilesional side, have revealed a higher prevalence after right hemisphere lesions [11,21,22]. Our results showed that the CED was specifically larger in stroke patients with spatial neglect than in stroke patients and in non-brain damaged subjects without the disorder. These results confirm earlier observations [7,8] in a newly recruited sample of subjects. Beyond, the present study now allowed for the first time to determine a cut-off threshold of 14.1° of horizontal eye-in-head deviation, allowing to differentiate right hemispheric stroke patients with versus without spatial neglect with a sensitivity of 96.4% and a specificity of 100%. This became possible by automatically playing the simple instruction “Please look straight ahead” whenever the program for a cranial CT was started. The instruction was first given when the patient was positioned on the CT-table and, a second time, immediately before the scan started. The beauty of this procedure was that the neuroradiology staff did not have to pay attention to and remember any changes in their normal routine. Such a simple addition can be part of the normal routine operations of any neuroradiologic unit without major staff briefings. It provides a new diagnostic tool for the dominant behavioral dysfunction after a right hemispheric lesion.

This new procedure allows the diagnosis of spatial neglect right at admission in the (hyper)acute phase of stroke i.e., long before any paper-and-pencil tests become available (in the present patient sample: 4 times earlier). This aspect opens a very significant new perspective for the treatment of spatial neglect. It now becomes theoretically possible to use specific therapies like, e.g., visual scanning training, active limb activation, or neck muscle vibration [23,24], already in a very early phase of the disease, i.e. before formal neuropsychological diagnostic testing can be applied. This can be the starting point to see whether an earlier diagnosis of spatial neglect can further increase the chance of its significant clinical improvement after right hemispheric stroke.

The routine stroke protocol in our Department carried out at admission composes the National Institutes of Health Stroke Scale (NIH-SS) [25]. Item 11 of the scale evaluates extinction and inattention, searching for signs of a spatial neglect [14]. Interestingly, only 41,7% of our patients with spatial neglect and 12,5% of our patients without spatial neglect (confirmed through the detailed neuropsychological paper pencil tests [see methods chapter above]) were recognized as having possible spatial neglect by item 11 of the scale. It is important to note that the initial NIH-SS was performed by the respective neurologist on duty and was not particularly supervised. Specific training and standardization to perform item 11 of the NIH-SS probably would increase the detection rate. Nevertheless, the low detection rate in the current sample obtained in a normal clinical setting demonstrates the need for an early, sensitive, and standardized method to verify the presence of spatial neglect on admission. The simple addition of the verbal command “Please look straight ahead” to the radiological routine procedure could provide such a tool.

The regular implementation of the verbal command in the scanner program for a regular cranial CT does not seem to have negative effects. This is also true for those CT programs that specifically try to avoid x-ray radiation of the lens or use eye-lens shielding for protection [23-26]. In the latter case, the verbal command “Please look straight ahead” would simply be useless, as eye deviation would not be measured afterwards, but would not delay or interfer with data aquisition. In general, it can be stated that - taking the average age on stroke patients [30] and the average dose of radiation to the unprotected lens per CT scan [31] in consideration - there seems to be a very small risk of developing cataracts due to cumulation of radiation, even in younger patients [31]. Thus, in our opinion the benefit of an early diagnosis of spatial neglect clearly overcome this specific risk. Nevertheless, the advantages and disadvantages of eye-lens shielding should be carefully weighed for each individual patient.

Conjugate eye deviation with an angle larger than 14° to the ipsilesional side has previously been shown to be a diagnostic predictor of acute and subacute ischemic stroke on supratentorial regions [32]. However, this latter study did not test whether the subjects with CED larger than 14° were subjects suffering from spatial neglect. In line with earlier work [7], the present investigation revealed that a horizontal CED larger than 14.1° is specifically associated with spatial neglect after right brain damage rather than with right brain damage per se.

It is important to note that the deviation value of 14.1° to the ipsilesional side resulting from the present work, was determined in a well selected sample of patients, namely patients with a first-time right hemisphere stroke who were able to perform the neuropsychological paper-pencil tests in an accurate matter and had no previous history of known cognitive impairment. The main goal of such a restricted group was to ensure that the patients studied could reliably focus on the verbal command. Variables that could cause inattention to the command itself should be largely excluded in order to assess the direct effect of the verbal command on the patients’ eye position. Despite this restriction in the present study, we expect that the very simple command “Please look straight ahead” also is possible to be executed by many subjects with cognitive decline. But still, of course, this matter needs further investigation in future studies, including a more heterogeneous sample of acute stroke patients. Beyond, it is also important to remember that the present procedure was not validated on patients with left brain damage. Due to the expected rate of disturbances of language prehension in these patients [10], it is possible to find larger variation of the CED value in this group. Another point to consider is that anatomical landmarks used to measure CED according to the procedure described by Simon et al. [11] have an interindividual variation [33–36]. This could make the cut-off deviation value of 14.1° appear to be examiner-dependent, at least to some degree. Future blinded studies with multiple investigators will help to adjust this possible variation.

To conclude, measuring the horizontal deviation of eye-in-head position on routine CT scans if subjects are instructed to “look straight ahead” during scanning, provides a promising new tool for early diagnosis of spatial neglect right at admission. It allows the therapeutic team to begin a deficit-oriented therapy early after stroke-onset to reduce hospitalization time and improve functional recovery.

## Data Availability

All data produced in the present study are available upon reasonable request to the authors.

## Acknowledgment

This work was supported by the Deutsche Forschungsgemeinschaft (KA 1258/23-1). We thank Daniel Wiesen for his help with the clinical neuropsychological training of JCM, Lisa Röhrig for her help with the lesion analysis, and Professors Martin Lenz and Stephan Clasen from the Department of Diagnostic and Interventional Radiology at Kreiskliniken Reutlingen GmbH for their consent to conduct the investigation.

